## Supplementary figures and images for "Applying GAN-based data augmentation to improve transcriptome-based prognostication in breast cancer"

### Supplementary Fig. 1

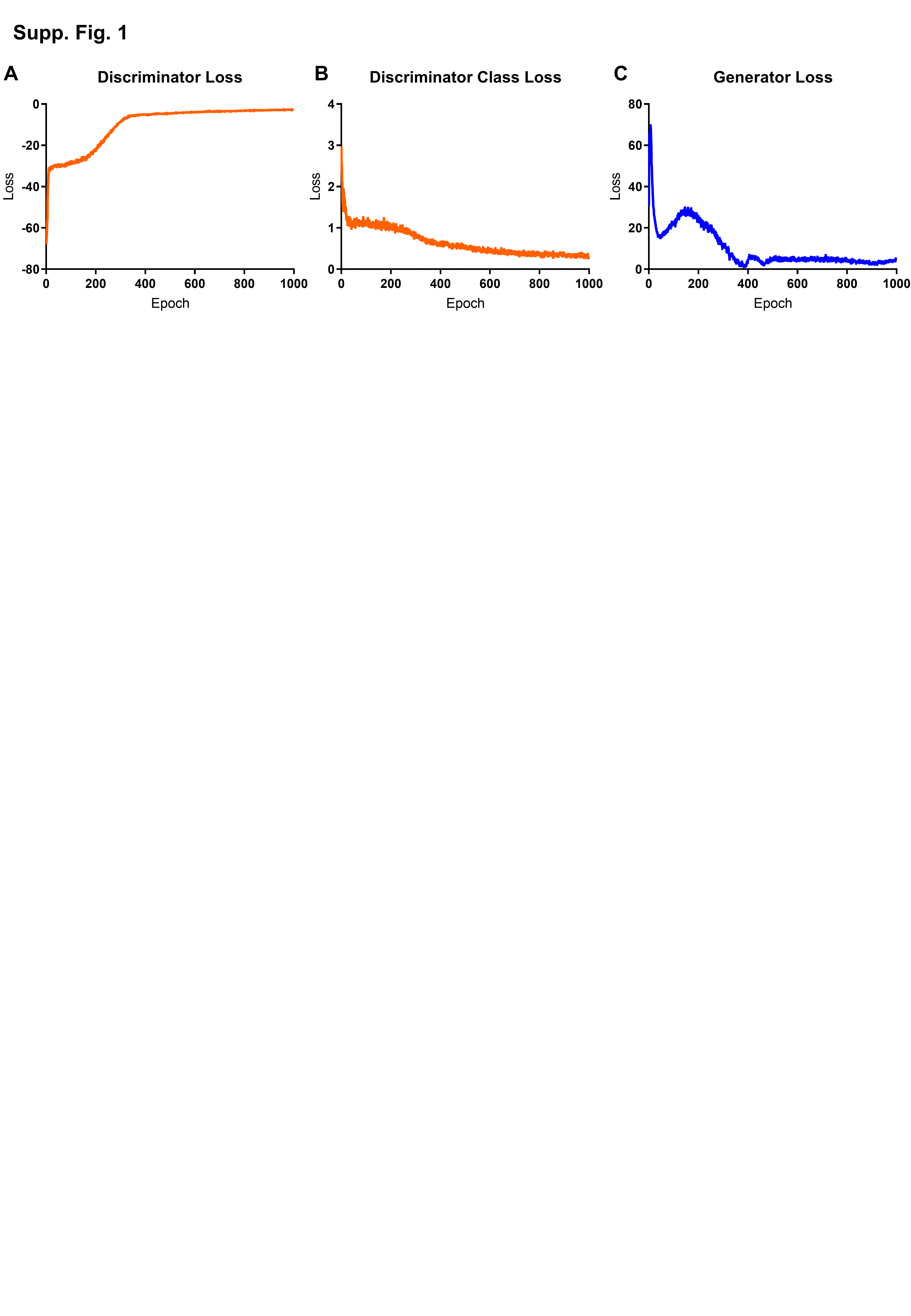

### Supplementary Fig. 2

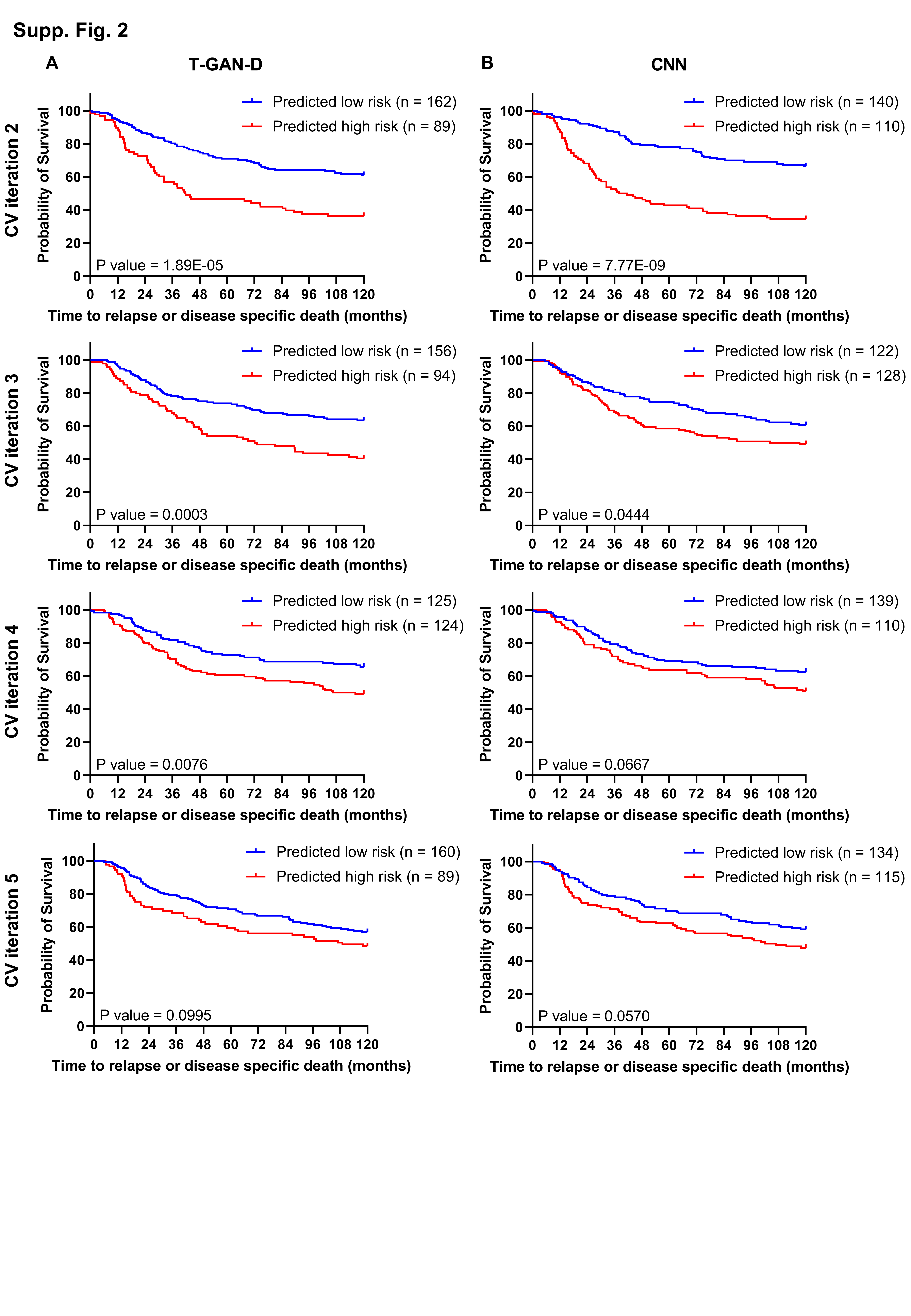

### Supplementary Fig. 3

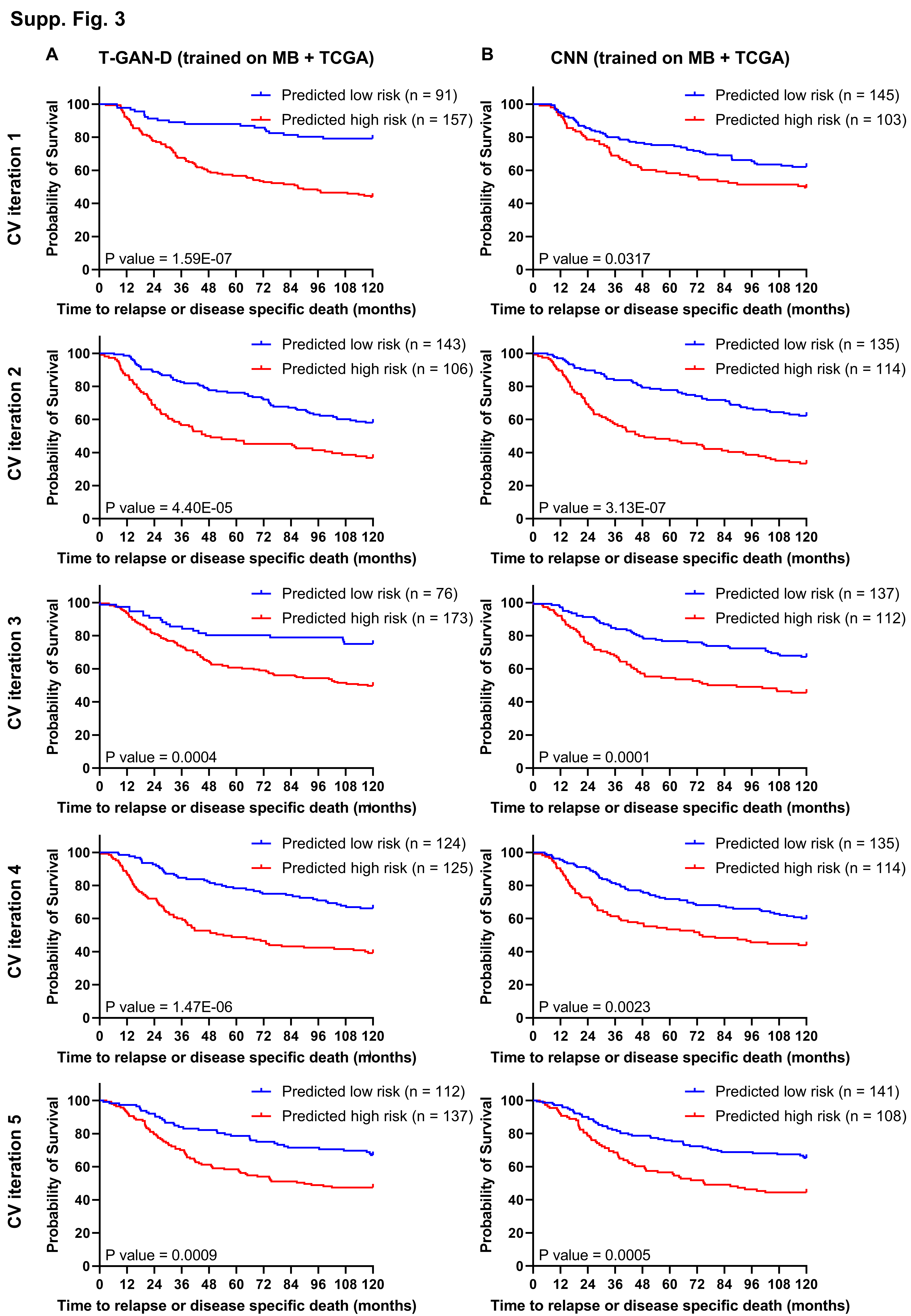

### Supplementary Fig. 4

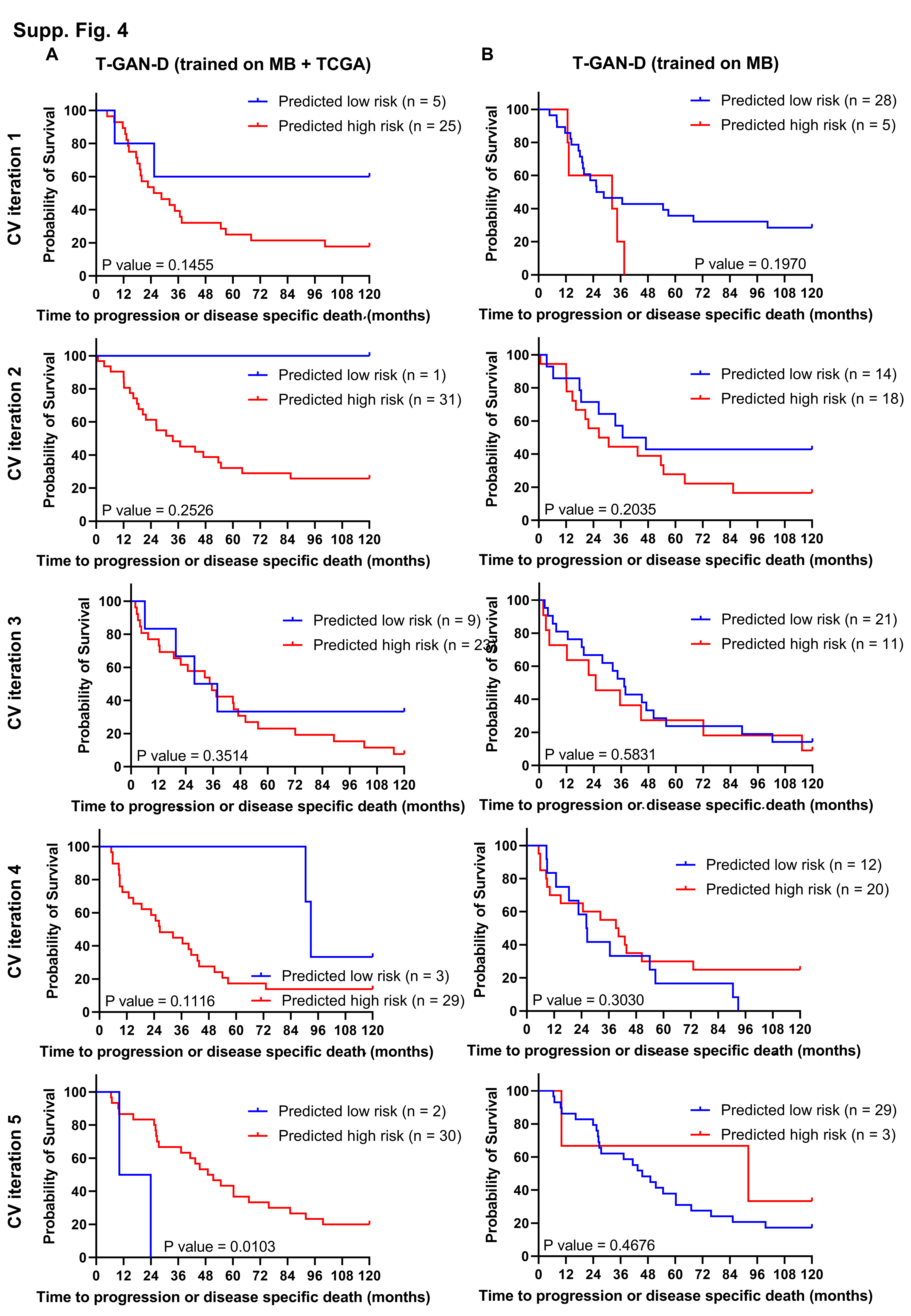
