## Supplementary tables for "Applying GAN-based data augmentation to improve transcriptome-based prognostication in breast cancer"

| **Supplementary Table 1** |  | |  | |
| --- | --- | --- | --- | --- |
| **MB risk class prediction**  **(Fig. 2, Supp Fig. 2)** | **T-GAN-D** | | **CNN** | |
| **Run no.** | **Accuracy** | **Log-rank P value** | **Accuracy** | **Log-rank P value** |
| **1** | 61.2% | 6.96E-05 | 55.2% | 0.1424 |
| **2** | 62.0% | 1.89E-05 | 66.0% | 7.77E-09 |
| **3** | 62.0% | 0.0003 | 55.6% | 0.0444 |
| **4** | 58.2% | 0.0076 | 56.6% | 0.0667 |
| **5** | 55.0%% | 0.0995 | 55.8% | 0.0570 |
| **Mean accuracy and pooled Log-Rank P value** | **59.7%** | **2.71E-12** | **57.9%** | **9.41E-09** |
| **Supplementary Table 2** |  |  |  |  |
| **MB risk class prediction**  **(Fig. 3, Supp. Fig. 3)** | **T-GAN-D  (trained on MB + TCGA)** | | **CNN**  **(trained on MB + TCGA)** | |
| **Run no.** | **Accuracy** | **Log-rank P value** | **Accuracy** | **Log-rank P value** |
| **1** | 64.5% | 1.59E-07 | 57.3% | 0.0317 |
| **2** | 60.2% | 4.40E-05 | 64.3% | 3.13E-07 |
| **3** | 57.8% | 0.0004 | 61.4% | 0.0001 |
| **4** | 63.5% | 1.47E-06 | 58.2% | 0.0023 |
| **5** | 62.7% | 0.0009 | 61.0% | 0.0005 |
| **Mean accuracy and pooled Log-Rank P value** | **61.7%** | **< 1E-15** | **60.5%** | **< 1E-15** |
| **Supplementary Table 3** |  |  |  |  |
| **TCGA risk class prediction**  **(Fig. 5, Supp Fig. 4)** | **T-GAN-D  (trained on MB + TCGA)** | | **T-GAN-D  (trained on MB)** | |
| **Run no.** | **Accuracy** | **Log-rank P value** | **Accuracy** | **Log-rank P value** |
| **1** | 78.8% | 0.1455 | 39.4% | 0.1970 |
| **2** | 75.0% | 0.2526 | 65.6% | 0.2035 |
| **3** | 81.3% | 0.3514 | 40.6% | 0.5831 |
| **4** | 81.3% | 0.1116 | 46.9% | 0.3030 |
| **5** | 75.0% | 0.0103 | 21.9% | 0.4676 |
| **Mean accuracy and pooled Log-Rank P value** | **78.3%** | **0.0623** | **42.9%** | **0.3497** |
