## Supplementary figure legends for "Applying GAN-based data augmentation to improve transcriptome-based prognostication in breast cancer"

**Supp. Fig. 1:** AC-WGAN-GP loss functions.

**(A)** Loss functions of the discriminator identifying real vs fake patients and **(B)** risk category. **(C)** Loss function of the generator. Loss functions were computed over 1000 training epochs.

**Supp. Fig. 2:** Kaplan-Meier curves generated with the risk categories predicted in the CV iterations not shown in Fig. 2. The prototyping MB cohort with all available transcriptomic data was used to compare the patient stratification obtained with the (**A**) T-GAN-D and (**B**) a classic CNN.

**Supp. Fig. 3:** Kaplan-Meier curves of individual CV iterations pooled in Fig. 3. A fraction of the MB and the full TCGA cohorts were integrated to train (**A**) the T-GAN-D and (**B**) the CNN. After rescaling both datasets and filtering out the genes not available in both cohorts the risk class of the MB patients was predicted.

**Supp. Fig. 4:** Kaplan-Meier curves of individual CV iterations pooled in Fig. 5. The T-GAN-D was trained (**A**) on the merged dataset and (**B**) on the MB dataset alone. After rescaling both datasets and filtering out the genes not available in both cohorts the risk class of the TCGA patients was predicted.

**Supp. Table 1-3: Accuracy and Log-rank P value of each CV iteration and pooled category predictions for all experimental settings.**
